# Pre-pandemic cognitive function and COVID-19 mortality: prospective cohort study

**DOI:** 10.1101/2021.02.07.21251082

**Authors:** G. David Batty, Ian J. Deary, Catharine R. Gale

**Author notes:** Correspondence: David Batty, Department of Epidemiology & Public Health, University College London, 1-19 Torrington Place, London, UK, WC1E 6BT.

## Abstract

**Background:** Poorer performance on standard tests of cognitive function is related to an elevated risk of death from lower respiratory tract infections. Whether pre-pandemic measures of cognition are related to COVID-19 mortality is untested.

**Methods:** UK Biobank, a prospective cohort study, comprises around half a million people who were aged 40 to 69 years at study induction between 2006 and 2010 when a reaction time test was administered to the full sample, and verbal-numeric reasoning assessed in a subgroup. Death from COVID-19 was ascertained from participant linkage to a UK-wide national registry.

**Results:** Between April 1^st^ and September 23^rd^ 2020, there were 388 deaths (138 women) ascribed to COVID-19 in the 494,932 individuals (269,602 women) with a reaction time test result, and 125 such deaths (38 women) in the 180,198 (97,794 women) for whom there were data on verbal-numeric reasoning. In analyses adjusted for age, sex, and ethnicity, a one standard deviation (118.2 msec) slower reaction time was related to a higher rate of death from COVID-19 (hazard ratio; 95% confidence interval: 1.18; 1.09, 1.28). A one standard deviation disadvantage (2.16 point) on the verbal-numeric reasoning test was also associated with an elevated risk of death (1.32; 1.09, 1.59). Attenuation after adjustment for additional covariates followed a similar pattern for both measures of cognition. For verbal-numeric reasoning, for instance, the hazard ratios were 1.22 (0.98, 1.51) after control for socioeconomic status, 1.16 (0.96, 1.41) after lifestyle factors, 1.25 (1.04, 1.52) after co-morbidity, and 1.29 (1.01, 1.64) after physiological indices.

**Conclusions:** In the present study, poorer performance on two pre-pandemic indicators of cognitive function, including reaction time, a knowledge-reduced measure, was related to death ascribed to COVID-19.

## Introduction

Cognitive function – also referred to as mental ability or intelligence – refers to psychological functions that involve the storage, selection, manipulation, and organisation of information, and the planning of actions. Evidence from well-characterised cohort studies suggest that lower scores on standard tests of cognition are linked to elevated rates of premature mortality,^1-4^ coronary heart disease,^5-9^ stroke,^7,10,11^ and intentional^12,13^ and unintentional injury.^14-17^ The magnitude of these effect estimates range between 2 and 6, and, while somewhat attenuated after control for covariates which include socioeconomic status and lifestyle factors, the gradients largely remain.

Similar inverse relations to those apparent for non-communicable disease and injury have recently been reported for cognition and death from respiratory infection, comprising pneumonia and influenza.^18^ Various explanations have been advanced, including the observation that people with higher cognitive ability are better equipped to obtain, process, and respond to disease prevention advice,^19^ as well as having healthier behaviours which include a lower prevalence of cigarette smoking,^20,21^ itself a risk factor for pneumonia.^22^ Additionally, people with higher educational achievement—strongly related to cognition^23^— are more likely to take up influenza and pneumococcal vaccination.^24^ These observations recently led us to examine if cognitive function in adults was related to the risk of hospitalisation for COVID-19. Of a range of baseline psychosocial factors including mental health, socioeconomic status, and personality disposition, cognitive function was the most strongly predictive of hospitalisation for COVID-19, whereby a doubling of disease risk was apparent in the lowest scoring group.^25^ With the pandemic evolving since we published those analyses using UK Biobank, there is now a sufficiently high number of deaths to examine if similar results are apparent for pre-pandemic cognitive function and COVID-19 mortality – an association which, to the best of our knowledge, has yet to be tested.

## Methods

Between 2006 to 2010, baseline data were collected in the UK Biobank, a prospective cohort study. Conducted across 22 research assessment centres, a total of 502,655 individuals aged 40 to 69 years participated (response 5.5%).^26,27^ Ethical approvals were received from the North-West Multicentre Research Ethics Committee. Data are publicly available upon application (https://www.ukbiobank.ac.uk/).

### Assessment of cognitive function

We use two tests of cognitive function as administered at baseline. Verbal and numeric (‘verbal-numeric’) reasoning was assessed using a computerized 13-item multiple-choice test with a two-minute time limit. The score derived was the number of correct answers. This test was introduced during the baseline assessment period; therefore, data are available for a subset of study members only (N=180,914). Reaction time, which is thought to capture efficiency of speed of information processing, is a knowledge-reduced indicator of cognition. Measured in the present study using a computerized Go/No-Go “Snap”-type game (N=496,882), participants were presented with electronic images of two cards. If symbols on the cards were identical, participants were instructed to immediately push the button-box using their dominant hand. The first five pairs were used as a practice with the remaining seven pairs, containing four identical cards, forming the assessment. Reaction time score was the mean time (milliseconds) taken to depress the button after each of these four matching pairs was presented. Reaction time correlates with cognitive tests that involve complex reasoning and knowledge such that people with higher cognitive ability tend to have faster reaction times.^28^ Scores from both tests show moderately high correlations with other measures of cognition and 4-week test-rest stability is high.^29^

### Assessment of confounding factors

Covariates were also assessed at baseline. Socioeconomic status was quantified using self-reported educational qualifications (degree, other qualifications, no qualifications), occupational classification based on current job, and the Townsend index of neighbourhood deprivation (higher scores denote greater disadvantage). Ethnicity was categorised as White, Asian, Black, Chinese, Mixed, or ‘other’ ethnic group. Vascular or heart problems, diabetes, chronic bronchitis or emphysema, and asthma, were based on self-report of a physician diagnosis. Hypertension was defined as systolic/diastolic blood pressure ≥ 140/90 mmHg and/or self-reported use of antihypertensive medication. Study members were also asked whether they had ever been under the care of a psychiatrist for any mental health problem.^30^ C-reactive protein, glycated haemoglobin, and high-density lipoprotein cholesterol concentrations were based on assays of non-fasting venous blood. Height, weight–from which body mass index was computed–and forced expiratory volume in one second were measured using standard protocols. Cigarette smoking, physical activity, and alcohol consumption were assessed using standard enquiries. Study members were linked to national mortality records and death from COVID-19, our outcome of interest, was denoted by the emergency ICD-10 code U07.1 (COVID-19, virus identified).

### Statistical analyses

To summarise the relation between cognition and mortality we used Cox regression to compute hazard ratios and with accompanying 95% confidence intervals.^31^ In these analyses we calculated effect estimates for tertiles of scores for both the test of verbal-numeric reasoning (<4 [most disadvantaged], 5-6, ≥7) and reaction time (≤499 msec, 500-581, ≥582 [most disadvantaged]). We also computed hazard ratios for a unit (standard deviation) disadvantage in score for verbal numeric reasoning (per 2.16 point decrease) and for reaction time (per 118.2 msec increase). The most basic analyses were adjusted for known COVID-19 risk factors (age, sex, and ethnicity^25,32,33^). Retaining these basic covariates, we then explored the impact of separately controlling for socioeconomic circumstances, existing medical conditions, lifestyle factors, and biological indices.

## Results

In the analytical sample of 494,932 individuals (269,602 women) with complete data on reaction time plus basic covariates (age, sex, ethnicity), 388 deaths (138 in women) were ascribed to COVID-19 between April 1^st^ and September 23^rd^ 2020. In the subgroup with data on the test of verbal-numeric reasoning, there were 125 such deaths (38 in women) in 180,198 people (97,794 women).

In table 1, in age-, sex- and ethnicity-adjusted analyses, we show, individually, the relation of the covariates and cognitive exposures with death from COVID-19. Twenty covariates were related to a higher risk of death from COVID-19 in these minimally-adjusted analyses; only the point estimate for regular intake of alcohol and asthma did not achieve statistical significance at conventional levels. Thus, there was a raised risk of COVID-19 death in people who were older, male, of ethnic minority ancestry, and socioeconomically disadvantaged. Those study members who smoked, reported less physical activity, and had higher body mass index, those with extant illness at baseline, and those with unfavourable levels of known cardiovascular disease biomarkers – lower lung function, higher systemic inflammation, lower high-density lipoprotein, and higher glycated haemoglobin – also experienced elevated rates of COVID-19 mortality.

**Table 1.** Cognitive function and covariates at baseline (2006-2010) according to death from COVID-19 (2020)

|  | COVID-19 mortality |  | P value | Hazard ratios (95% CI) |
| --- | --- | --- | --- | --- |
|  | Yes (n=404) | No (n=502,251) |  |  |
| *Demographic factors* |  |  |  |  |
| Age, yr, mean (SD) | 62.7 (6.03) | 56.5 (8.09) | <0.0001 | 3.07 (2.67, 3.53) |
| Female, N (%) | 145 (35.9) | 273,327 (54.4) | <0.0001 | 0.48 (0.38, 0.59) |
| Non-white ethnicity | 37 (9.27) | 26,997 (5.41) | 0.001 | 3.75 (2.67, 5.27) |
| *Comorbidities* |  |  |  |  |
| Vascular or heart disease, N (%) | 216 (54.3) | 149,139 (29.8) | <0.0001 | 1.79 (1.46, 2.19) |
| Hypertension, N (%) | 313 (79.9) | 282,324 (57.2) | <0.0001 | 1.70 (1.32, 2.19) |
| Diabetes, N (%) | 63 (15.8) | 26,345 (5.27) | <0.0001 | 2.31 (1.75, 3.06) |
| Chronic bronchitis or emphysema, N (%) | 18 (4.46) | 8,334 (1.66) | <0.0001 | 2.37 (1.47, 3.80) |
| Asthma, N (%) | 34 (8.42) | 57,879 (11.5) | 0.050 | 0.80 (0.56, 1.14) |
| Mental health - Psychiatric consultation, N (%) | 56 (14.1) | 57,625 (11.6) | 0.117 | 1.42 (1.07, 1.88) |
| *Lifestyle factors* |  |  |  |  |
| Current smoker, N (%) | 50 (12.6) | 52,580 (10.5) | <0.0001 | 1.88 (1.37, 2.59) |
| No physical activity, N (%) | 55 (14.0) | 32,804 (6.63) | <0.0001 | 2.54 (1.90, 3.40) |
| Drinks alcohol daily/almost daily, N (%) | 85 (21.3) | 101,707 (20.3) | 0.641 | 0.88 (0.69, 1.12) |
| Body mass index, kg/m<sup>2</sup>, mean (SD) | 29.7 (5.83) | 27.4 (4.80) | <0.0001 | 1.51 (1.39, 1.65) |
| *Biomarkers* |  |  |  |  |
| Lung function, L, mean (SD) | 2.53 (0.87) | 2.81 (0.80) | <0.0001 | 0.69 (0.60, 0.80) |
| C-reactive protein, mg/L, median (IQR) | 1.72 (0.83, 3.51) | 1.26 (0.63, 2.49) | 0.0001 | 1.26 (1.12, 1.41) |
| High-density lipoprotein, mmol/L, median (IQR) | 1.26 (1.09, 1.52) | 1.40 (1.17, 1.67) | 0.0001 | 0.79 (0.69, 0.90) |
| HbA1C, mmol/mol, median (IQR) | 36.7 (33.8, 40.7) | 35.2 (32.8, 37.9) | 0.0001 | 1.25 (1.16, 1.35) |
| *Socioeconomic factors* |  |  |  |  |
| No university education, N (%) | 310 (80.7) | 330,988 (67.3) | <0.0001 | 1.75 (1.35, 2.26) |
| Neighbourhood deprivation score, median (IQR) | -0.887 (-3.04, -2.48) | -2.14 (-3.64, 0.55) | 0.0001 | 1.45 (1.33, 1.59) |
| Personal service, sales occupations etc, N (%) | 54 (27.8) | 66,160 (19.0) | 0.002 | 1.57 (1.15, 2.16) |
| *Psychological factors* |  |  |  |  |
| Verbal-numeric reasoning, mean (SD) | 5.32 (2.20) | 6.02 (2.16) | 0.0002 | 1.32 (1.09, 1.59) |
| Reaction time, msec, mean (SD) | 606.1 (136.4) | 559.9 (118.2) | <0.0001 | 1.18 (1.09, 1.28) |

Hazard ratios are adjusted for age, sex, and ethnicity and expressed per category, or per SD increase for continuous variables (except for reasoning which is expressed per SD decrease [disadvantage]). The maximum analytical sample of 502,655 people was lower in selected analyses owing to missing data.

In analyses adjusted for age, sex, and ethnicity, individuals with lower verbal-numeric reasoning scores had a higher risk of death ascribed to COVID-19 (hazard ratio per SD disadvantage; 95% confidence interval: 1.32; 1.09, 1.59) (tables 1 and 2). Whereas adjusting for markers of socioeconomic position and biological factors had little impact on these effect estimates, adding co-morbidity to the multivariable model led to some attenuation (1.25; 1.04, 1.52). The greatest degree of attenuation was evident after adjusting for lifestyle indices (1.16; 0.96, 1.41) which included smoking and body weight.

**Table 2.** Hazard ratios (95% confidence intervals) for the association of measures of baseline cognitive function (2006-2010) with death from COVID-19 (2020)

|  | Adjusted for age, sex, and ethnicity | Adjusted for age, sex, ethnicity, and comorbidity | Adjusted for age, sex, ethnicity, and markers of socioeconomic status | Adjusted for age, sex, ethnicity, and lifestyle factors | Adjusted for age, sex, ethnicity, and biological factors |
| --- | --- | --- | --- | --- | --- |
| **Verbal-numeric reasoning** |  |  |  |  |  |
| *Cases/Number at risk* | *125/180,198* | *123/177,361* | *90/145,975* | *119/177,545* | *77/126,190* |
| 1 (most disadvantaged) | 2.04 (1.30, 3.20) | 1.86 (1.18, 2.93) | 2.22 (1.26, 3.94) | 1.52 (0.95, 2.43) | 2.32 (1.30, 4.14) |
| 2 | 1.35 (0.86, 2.14) | 1.29 (0.81, 2.03) | 1.34 (0.78, 2.31) | 1.27 (0.80, 2.01) | 1.62 (0.90, 1.89) |
| 3 | 1.0 (reference) | 1.0 | 1.0 | 1.0 | 1.0 |
| P for trend | 0.002 | 0.007 | 0.006 | 0.082 | 0.004 |
| Per SD disadvantage | 1.32 (1.09, 1.59) | 1.25 (1.04, 1.52) | 1.31 (1.02, 1.67) | 1.16 (0.96, 1.41) | 1.29 (1.01, 1.64) |
| **Reaction time** |  |  |  |  |  |
| *Cases/Number at risk* | *388/494,932* | *378/483,785* | *182/339,977* | *373/484,832* | *250/352,096* |
| 1 | 1.0 (reference) | 1.0 | 1.0 | 1.0 | 1.0 |
| 2 | 0.97 (0.73, 1.30) | 0.92 (0.69, 1.24) | 1.01 (0.67, 1.54) | 0.94 (0.70, 1.27) | 0.90 (0.62, 1.30) |
| 3 (most disadvantaged) | 1.50 (1.15, 1.95) | 1.40 (1.07, 1.83) | 1.66 (1.13, 2.44) | 1.35 (1.03, 1.77) | 1.50 (1.08, 2.09) |
| P for trend | 0.001 | 0.003 | 0.004 | 0.009 | 0.003 |
| Per SD disadvantage | 1.18 (1.09, 1.28) | 1.16 (1.07, 1.27) | 1.21 (1.07, 1.37) | 1.15 (1.06, 1.25) | 1.19 (1.07, 1.91) |

Comorbidities: diagnoses of vascular or heart disease, diabetes, chronic bronchitis or emphysema, asthma, hypertension, and mental illness. Socioeconomic status (SES): educational attainment, occupational classification, and area deprivation. Lifestyle factors: body mass index, smoking status, alcohol intake, and physical activity. Biomarkers: Forced expiratory volume in the first second, and blood concentrations of C-reactive protein, glycated haemoglobin, and high-density lipoprotein.

The associations with COVID-19 mortality are additionally depicted by tertiles of the cognitive exposures in table 2. In minimally-adjusted analyses (age, sex, ethnicity), relative to the highest performing tertile of verbal-numeric reasoning, those people in the lowest-scoring group experienced a doubling in the rate of death from COVID-19. There was also evidence of a dose-response effect such that the intermediate cognition group experienced an intermediate level of risk (P for trend: 0.002). In these analyses, although rates of COVID-19 mortality were still associated with around a 50% increase risk in the lowest cognition-scoring group, statistical significance at conventional levels was lost as evidenced by the confidence intervals crossing unity.

Slower responders to the reaction time test had an elevated risk of death from the disease (1.18; 1.09, 1.28) (tables 1 and 2). In contrast to the analyses for verbal-numeric reasoning, there was less attenuation of estimates after controlling for the same covariates. Adjustment for lifestyle factors again resulted in the greatest attenuation of the cognition–mortality gradient but this was modest (1.15; 1.06, 1.25); statistical significance was retained, despite a smaller effect size, owing to the larger analytical sample in the reaction time analyses. Unlike the analyses of verbal-numeric reasoning, however, when the results were analysed by tertiles, there appeared to be a threshold effect: only people in the slowest reaction time tertile had elevated risk; the hazard ratio in the intermediate group approximated 1.

Lastly, given the known correlation for education and verbal-numeric reasoning (r=0.40, p<0.0001, N=178,908 in the present data), we conducted separate adjustment for education, one of three indicators of socioeconomic status used herein (table 1a, appendix). Controlling for education had a marginal attenuating effect on the strength of the cognition–COVID-19 death association, whereas the addition of the other socioeconomic factors of area deprivation and occupational classification led to positive confounding such that the magnitude of categorical effects increased somewhat. It is also the case, however, that, owing to some missing data for occupation, the apparent difference in strength of the association may at least partly reflect differences in sample size. With reaction time being a knowledge-reduced measure of cognition, its relationship with education (r=0.15, p<0.0001, N=487,993) was weaker than for verbal-numeric reasoning.

## Discussion

Our main finding was that, net of several covariates, poorer scores on two tests of cognition – verbal-numeric reasoning and reaction time – were associated with a higher risk of death from COVID-19. Patterns of attenuation were similar for both exposures such that the greater impact was apparent after control for lifestyle factors, whereas physiological indices had little if any attenuating effect. We also replicated known risk factors for death from COVID-19 – being older, male, from an ethnic minority, socioeconomically disadvantaged, and having an extant somatic medical condition have been repeatedly linked with poor prognosis in COVID-19 patients in the US,^32^ UK,^34^ Italy,^35^ China,^36,37^ and Brazil.^38^ The raised risk of COVID-19 mortality seen in people with the most disadvantaged scores for reaction time – as described, a knowledge-reduced measure of cognition – corroborates the results for verbal-numeric reasoning which is more closely linked to education. It is also the case that the magnitude of association for scores on both tests were little attenuated after taking into account educational attainment.

### Plausible mechanisms

A plausible explanation for the association between cognition and respiratory infection in general is that people with higher ability are more likely to take-up influenza and pneumococcal inoculation. Although people with a higher level of education, a close correlate of mental ability,^1,23^ are less likely to be vaccine hesitant,^39^ that the present data were collected prior to the commencement of any UK-wide distribution of a COVID-19 vaccine renders this explanation implausible. A more likely line of reasoning is the deluge of health advice in the current pandemic during a period when news outlets and social media platforms have never been more ubiquitous. Preventative information has ranged from the simple and practical to the complex, contradictory, false, and fradulent.^40-42^ In order to diminish their risk of the infection, the population has to acquire, synthesise, and deploy this information but the ability to do so seems to vary by levels of health literacy^43^ just as it may for its close correlate, cognitive function.^44^

### Comparison with existing studies

The present findings can be most directly compared with those from an earlier study of hospitalisations,^25^ where, after control for multiple covariates, lower verbal-numeric reasoning based on the same categorisation as used herein was associated with a doubling of risk. Unlike the present analyses featuring death as the outcome of interest, in that study^25^ the role of reaction time as a predictive factor for hospitalisation for COVID-19 was less clear. In those analyses we used a record of a positive in-patient test for COVID-19 as our outcome of interest. Whereas this was assumed to be an indicator of disease severity – only serious cases are hospitalised in the UK which operates under a single, national health service – it is nonetheless plausible that some of the cases were patients being treated for unrelated conditions who were asymptomatically positive for COVID-19 after routine hospital-wide testing. Our results here for death from the disease, not only for pre-pandemic cognitive function but also mental health problems,^30^ therefore corroborates those earlier findings.^25^

Some indirect support for the present results can be found in studies examining the association between dementia risk and COVID-19. Using a record linkage approach, individuals with pre-pandemic hospitalisation for dementia, in particular Alzheimer’s, experienced around a doubling of risk of later COVID-19 hospitalisation.^45^ In one of the largest studies to date – OpenSAFELY, comprising more than 17 million primary care-registered people – a record of either stroke and/or dementia was associated with around a doubling in the risk of a COVID-related death after multiple adjustment for other co-morbidities and ethnicity, but the separate impact of these neurological conditions was not reported.^46^

### Study strengths and weaknesses

The strengths of our study include the well-characterised nature of the sample and the full coverage of the population for cause of death. Our study has some weaknesses. Although the present cohort is large, only a subgroup was administered the verbal-numeric reasoning tests and the relatively modest number of deaths attributable to COVID-19 in that sample perhaps resulted in lower-than-optimal levels of statistical precision. The cognitive tests are brief—especially the reaction time test—and this is likely to reduce effect sizes of associations. Nevertheless, both cognitive tests do demonstrate some concurrent validity with longer and better-validated tests.^29^ With the present sample not being representative of the general UK population, death rates from leading causes and the prevalence of reported risk factors are known to be underestimates of those apparent in less select groups;^27^ the same is likely to be the case for COVID-19 deaths. This notwithstanding, there is evidence that, for major causes of death, risk factor associations are externally valid.^27^ Lastly, levels of our baseline data – exposures and covariates – will vary in the period between study induction in UK Biobank and the present pandemic. This is a perennial issue in cohort studies and one we were able to investigate using data from a resurvey that took place around 8 years after baseline examination in a sub-sample.^47^ Analyses revealed moderate-to-high stability for verbal-numeric reasoning (r=0.63, p<0.001, N=9689) and reaction time (r=0.49, p<0.001, N=28,810), and key covariates such as education (r=0.86, p<0.001, N=30,350) and co-morbidities such as diabetes (r=0.63, P<0.001, N= 31,037).

In conclusion, in the present study, poorer performance on two pre-pandemic indicators of cognitive function, including reaction time, a knowledge-reduced measure, was related to death ascribed to COVID-19.

## Data Availability

Data from UK Biobank (http://www.ukbiobank.ac.uk/) are available to bona fide researchers upon application.

## Acknowledgement

We thank UK Biobank study members for their generosity in participating.

## Access to data

Data from UK Biobank (http://www.ukbiobank.ac.uk/) are available to *bona fide* researchers upon application. Part of this research has been conducted using the UK Biobank Resource under Application 10279.

## Notes

**Funding:** GDB is supported by the UK Medical Research Council (MR/P023444/1) and the US National Institute on Aging (1R56AG052519-01; 1R01AG052519-01A1); and IJD by the UK Medical Research Council (MR/R024065/1), UK Economic and Social Research Council (ES/S015604/1), and US National Institute on Aging (NIH), US (1R01AG054628-01A1); These funders, who provided no direct financial or material support for the work, had no role in study design, data collection, data analysis, data interpretation, or report preparation.

### Competing Interest Statement

The authors have declared no competing interest.

### Funding Statement

GDB is supported by the UK Medical Research Council (MR/P023444/1) and the US National Institute on Aging (1R56AG052519-01; 1R01AG052519-01A1); and IJD by the UK Medical Research Council (MR/R024065/1), UK Economic and Social Research Council (ES/S015604/1), and US National Institute on Aging (NIH), US (1R01AG054628-01A1); These funders, who provided no direct financial or material support for the work, had no role in study design, data collection, data analysis, data interpretation, or report preparation.

### Author Declarations

Ethical approval for UK Biobank was granted to the principal investigators by the North-West Multi-centre Research Ethics Committee, and the research was carried out in accordance with the Declaration of Helsinki of the World Medical Association; participants gave written consent. The present analyses of existing, anonymised data from the study has been conducted using the UK Biobank Resource under Application number 10279.

