## Supplemental table 1 for "Pre-pandemic cognitive function and COVID-19 mortality: prospective cohort study"

**Table 1a. Hazard ratios (95% confidence intervals) for the association of measures of baseline cognitive function (2006-2010)**

**with death from COVID-19 (2020) – individual adjustment for education**

|  | **Adjusted for age, sex and ethnicity** | **Adjusted for age, sex, ethnicity and educational qualifications** | **Adjusted for age, sex, ethnicity, and markers of socioeconomic status** |
| --- | --- | --- | --- |
| **Verbal-numeric reasoning** |  |  |  |
| *Cases/Number at risk* | *125/180,198* | *120/178,247* | *90/145,975* |
| 1 (most disadvantaged) | 2.04 (1.30, 3.20) | 1.81 (1.10, 2.98) | 2.22 (1.26, 3.94) |
| 2 | 1.35 (0.86, 2.14) | 1.20 (0.74, 1.93) | 1.34 (0.78, 2.31) |
| 3 | 1.0 (reference) |  | 1.0 |
| P for trend | 0.002 | 0.019 | 0.006 |
| Per SD (2.16 point) disadvantage | 1.32 (1.09, 1.59) | 1.25 (1.01, 1.54) | 1.31 (1.02, 1.67) |
| **Reaction time** |  |  |  |
| *Cases/Number at risk* | *388/494,932* | *372/486,265* | *182/339,977* |
| 1 | 1.0 (reference) | 1.0 | 1.0 |
| 2 | 0.97 (0.73, 1.30) | 0.92 (0.69, 1.24) | 1.01 (0.67, 1.54) |
| 3 (most disadvantaged) | 1.50 (1.15, 1.95) | 1.41 (1.07, 1.84) | 1.66 (1.13, 2.44) |
| P for trend | 0.001 | 0.003 | 0.004 |
| Per SD (118.2 msec) disadvantage | 1.18 (1.09, 1.28) | 1.17 (1.07, 1.27) | 1.21 (1.07, 1.37) |

Socioeconomic status (SES): educational attainment, occupational classification, and area deprivation
